# Drinking water quality and incidence of Diarrhoea in children aged 6 months to 15 years: Findings from a paediatric cohort in Vellore, Southern India

**DOI:** 10.64898/2026.01.13.26344065

**Authors:** Manikandan Srinivasan, S Vignesh Rajan, G Santhosh Kumar, N Samarasimha Reddy, Kulandaipalayam Natarajan Sindhu, Karthikeyan Ramanujam, Sathyapriya Subramaniam, Gagandeep Kang, Jacob John

## Abstract

**Introduction:** The coverage of access to basic drinking water and sanitation facilities in India was estimated to be 93% and 60%, respectively, in 2017. The monitoring of the burden of diarrhoeal illnesses, especially in children, remains important to assess the impact of the expansion of water and sanitation (WaSH) in the Indian setting. This study aimed to estimate the burden of diarrhoea in an established longitudinal pediatric cohort in an urban settlement of Vellore in South India.

**Methods:** The ‘Surveillance for Enteric Fever in India (SEFI)’ cohort established in an urban settlement of Vellore, south India, enrolled 6760 children aged between 6 months and 15 years. The cohort was followed up for typhoid and paratyphoid fever between 2017 and 2019. Field research assistants contacted caregivers of these children weekly to elicit any diarrhoeal illness in the child in the preceding week. As a part of SEFI environmental surveillance, drinking water samples from the study households were tested for coliforms. Sociodemographic characteristics, including source of drinking water, sanitation and hygiene practices, were collected. Incidence of diarrhoea was estimated and expressed as the number of diarrhoeal episodes over child-years of observation (CYO). Poisson regression analysis was performed to identify predictors of diarrhoeal episodes.

**Results:** The estimated incidence of diarrhoea in the 6501 children followed up between November 1, 2017, and October 31, 2019, was 31.1 episodes per 100 CYO, with children in the age group of 6 months and <5 years having a higher incidence of diarrhoea than those aged between 5 and 15 years (58.6 versus 22 episodes per 100 CYO). Of the 6467 children with information on WaSH available, 5812 (89.9%) used the public distribution system for drinking water. Of the 1804 drinking water samples tested, 1346 (74.6%) had coliform counts >10,000/100 mL. Only about one-third of the cohort (n=2293, 35.5%) lived in households with access to improved sanitation. Multivariable analysis showed that children aged <2 years, residing in crowded settlements, using the public distribution system for drinking water and from households with poor hygiene practices related to excreta disposal of under-five children had a higher risk for diarrhoea.

**Conclusion:** Approximately 8 in 10 children in urban Vellore lack access to safely managed drinking water, and thereby, are at a high risk for diarrheal illnesses, especially in the under-5 children. With rapidly expanding urbanisation in the Indian setting, it is pertinent that emphasis be laid on robust planning and provision of safely managed water and sanitation.

## Introduction

As per the global burden of diarrhoeal disease (GBD) estimates in 2021, diarrhoea was the leading cause of death among children aged <5 years, accounting for 340,000 deaths, with 58,584 (17.2%) reported from India [1]. Inclusive of the long-term sequelae of growth faltering due to infections, diarrhoeal diseases resulted in total DALYs of 39% [95% UI: 28.6 – 43.9] in under-5 children in India [2]. As per NFHS-4 estimates, the prevalence of diarrhoea among children under-5 years of age in Tamil Nadu was 8% in 2015-2016, with urban areas reporting a higher rate, 8.2%, compared to rural areas, 7.8% [3]. Cohort studies from India reported the incidence of diarrhoea in the under-5 age group ranging between 120 - 650 episodes per 100 child-years of observation (CYO), with children in urban areas having a two-fold increase in risk of diarrhoea compared to those from rural settings [4,5]. Information on the burden of diarrhoeal disease in older children (5–15 years) remains limited.

Environmental factors play a pivotal role in determining diarrhoeal disease transmission. Unsafe water sources and poor sanitation have been attributed to 72% (34% – 91.4%) and 56.4% (49.3% – 62.7%) diarrhoeal deaths among under-5 children globally [6]. A 10-fold increase in total coliform counts in drinking water samples has been implicated in four times greater rise of diarrhoeal cases in communities [7]. With regard to seasonal patterns over the past decade in India in diarrhoeal cases and deaths attributable to diarrhoea in children, smaller peaks of cases were observed during the summer (May), with a larger peak being reported during the monsoon (July) [8,9]. Given that scaling up access to improved sources of drinking water and sanitation, and basic hand washing practices are integral in diarrhoeal prevention, monitoring this improvement in access and practices in communities is important to reducing the diarrhoeal disease burden in India [10–12]. Thus, having reliable and context-specific data on the burden of diarrhoeal diseases and its risk factors periodically is crucial for understanding diarrhoeal disease epidemiology, enabling the tailoring of appropriate control strategies in India. Availability of longitudinal, intense surveillance mechanisms to capture information on diarrhoea cases, quality of drinking water and WaSH practices in the urban community of Vellore in south India as a part of ‘Surveillance for enteric fever in India’ (SEFI) paediatric cohort provided an opportunity to estimate diarrhoeal burden in this cohort and to examine the risk factors attributable to diarrhoea.

## Methods

The SEFI Vellore cohort was established in 2017 in an urban settlement of Vellore town in Tamil Nadu, a southern state in India. The cohort recruited about 6000 children from four contiguous urban settlements, namely Chinallapuram (CAP), Ramanaickanpalayam (RNP), Kaspa and Vasanthapuram (VSPM) and followed-up between November 1, 2017, and October 31, 2019 [13]. Study participants were recruited after obtaining a written informed consent [13]. This population depends on daily wage work for their livelihood, with beedi rolling (indigenous cigarette made with unprocessed tobacco) being their predominant occupation. The study population had access to healthcare services delivered through Christian Medical College and Hospital, a referral not-for-profit organisation, its satellite primary and secondary care hospitals, and state-run health centres and hospitals within a few kilometres of the study area. The study population also availed health services from private nursing homes and general practitioners in the area [14].

Children in this longitudinal community-based cohort were actively followed up for two years, or till their 15^th^ birthday, through an intense weekly surveillance for fever, diarrhoea and other common childhood illnesses. Primarily, this cohort was established to estimate the incidence of blood culture-confirmed typhoid fever in children. Trained field research assistants (FRAs) made weekly home visits to interview the primary caregivers of the children and documented any episode of diarrhoea (3 or more episodes of stools with altered consistency in a day) in the last seven days. Phone call-based surveillance was attempted if the primary caregivers were unavailable during home visits. FRAs completed weekly surveillance forms using electronic tablets via an in-house developed android application. Weekly surveillance visits were considered valid if the forms were completed within a maximum recall period of 14 days from the actual date of the visit. To ensure quality control of the weekly surveillance data, a part of the weekly surveillance data forms completed by the FRAs was validated by the field supervisors, where the supervisors completed the validation through a home visit or a phone surveillance [14].

Annual surveys were conducted to capture information on sociodemographic characteristics, source of drinking water, water treatment practices, sanitation facilities accessed by the household, and hygiene practices at study households using a standardised questionnaire [15]. Sources of drinking water, such as public distribution system and water supplied by mobile tankers, were classified as ‘unimproved sources’, while borewell and bottled water sources were classified as ‘improved sources’[16]. Regarding sanitation facilities, households with toilet effluents being discharged into a septic tank were categorised as having ‘improved sanitation’, while those households with toilets that discharge their effluents into drains/elsewhere in the surroundings were classified as ‘unimproved sanitation’[16]. Food practices, including the consumption of ready-to-eat foods in the households, were elicited by asking if they buy breakfast/lunch/dinner from street vendors.

This community cohort also incorporated a surveillance for testing the quality of drinking water through samples collected from the study households during the study period. Using a stratified random sampling approach with replacement, households were selected for water sampling, where the sampling strata were the study area and surveillance months. Water samples were transported to the laboratory for testing within 4 hours from the point of collection, and were tested for total coliform counts (colony forming units or CFU/100 mL) and residual chlorine levels using standard methodology described elsewhere [7].

## Statistical analysis

Sociodemographic characteristics and total coliform positivity in water samples were summarised using percentages. Seasonal distribution of diarrhoea cases and coliform contamination across the four study areas during the surveillance period were represented using an area graph. Diarrhoeal episodes were summarised as count data. The incidence rate of diarrhoea was calculated as a fraction of the total number of diarrhoeal episodes reported during the surveillance period over child-years of observation (CYO) in the cohort. Since the weekly surveillance captured incident diarrhoeal episodes, in case of children reporting two or more diarrhoeal episodes in consecutive surveillance weeks, the incident diarrhoeal episode reported during the first week was considered as one episode of diarrhoea, and this was used for calculating the number of episodes. The total number of valid weekly visits was converted into CYO. A Poisson regression model was used to identify factors associated with the incidence of diarrhoea, and incidence risk ratios along with 95% confidence intervals, were reported. P-value <0.05 was considered statistically significant. Statistical analysis was conducted using STATA version 14 (Stata Statistical Software: Release 14, StataCorp LP, College Station, TX).

## Results

This paediatric cohort of 6501 children was actively followed up between November 1, 2017, and October 31, 2019, accumulating 11600.7 CYO (**Table 1**). About 3983 (61.3%) children in the cohort were between 5 and 15 years, with nearly equal representation of males and females. Over two-thirds of children, 4530 (70.1%) were from households with lower socioeconomic status, and 3910 (60.5%) lived in a nuclear family. With regard to the study area, 2086 (32.1%) children of the cohort resided in Kaspa, with 976 (15%) children belonging to the VSPM area. The RNP and Kaspa areas were densely populated urban settlements with adjoining houses without proper sanitation facilities and this in contrast to the CAP area that had houses relatively widely dispersed. Majority, 5812 (89.9%) children were from households that depended on unimproved drinking water sources, such as public distribution systems or water supplied by mobile tankers. Nearly 3702 (57.2%) children lived in households with unimproved sanitary facilities, with effluents from their toilets connected to an open drain nearby or elsewhere. One-fourth of children, 1596 (24.7%), resided in households that practised disposal of their children’s stools into the drains around the houses or in garbage (**Table 2**).

**Table 1:** Incidence rate of diarrhoea per 100 child-years of observation (CYO) among children aged 6 months to 15 years of SEFI cohort in Vellore, India, 2017-19 (N=6501)

|  | No. of diarrhoea episodes | CYO | Incidence per 100 CYO |
| --- | --- | --- | --- |
| Overall incidence of diarrhoea | 3612 | 11600.7 | 31.1 (30.1 – 32.2) |
| Age-stratified incidence of diarrhoea (0.5 – 5 years) | 1696 | 2895.3 | 58.6 (55.8 – 61.4) |
| Age-stratified incidence of diarrhoea (5 – 15 years) | 1916 | 8705.4 | 22 (21 – 23) |

**Table 2:**
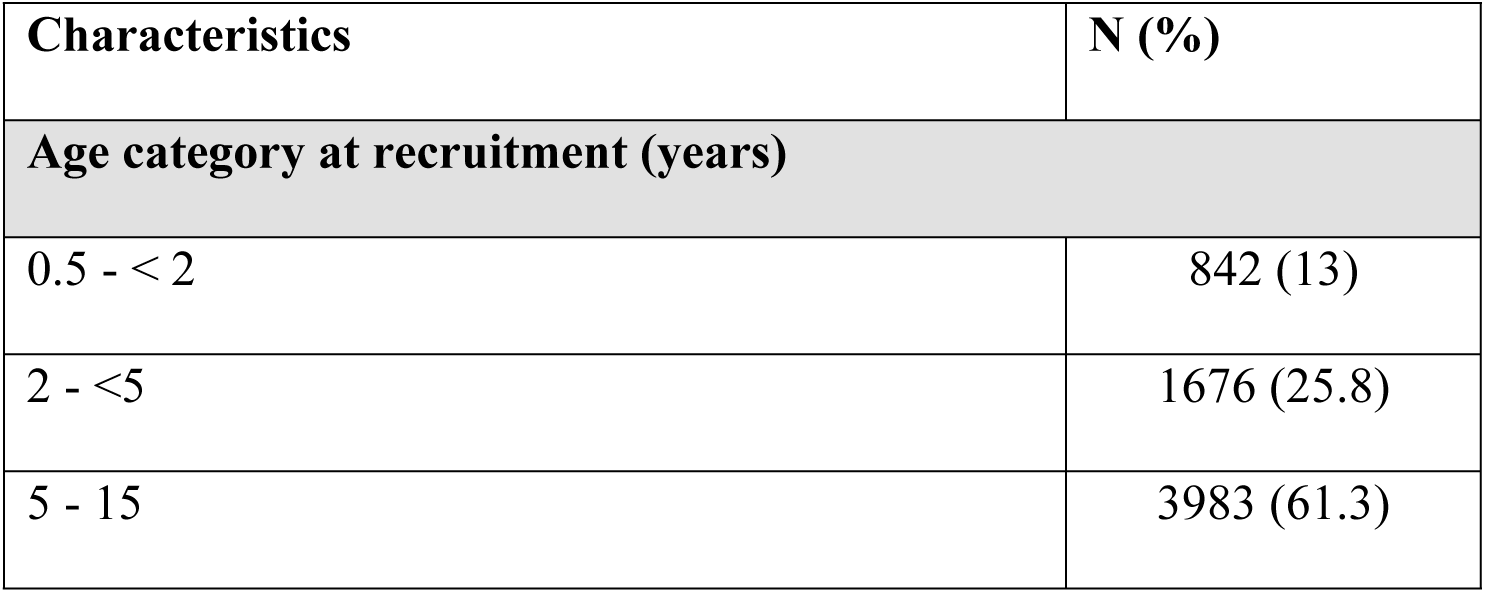
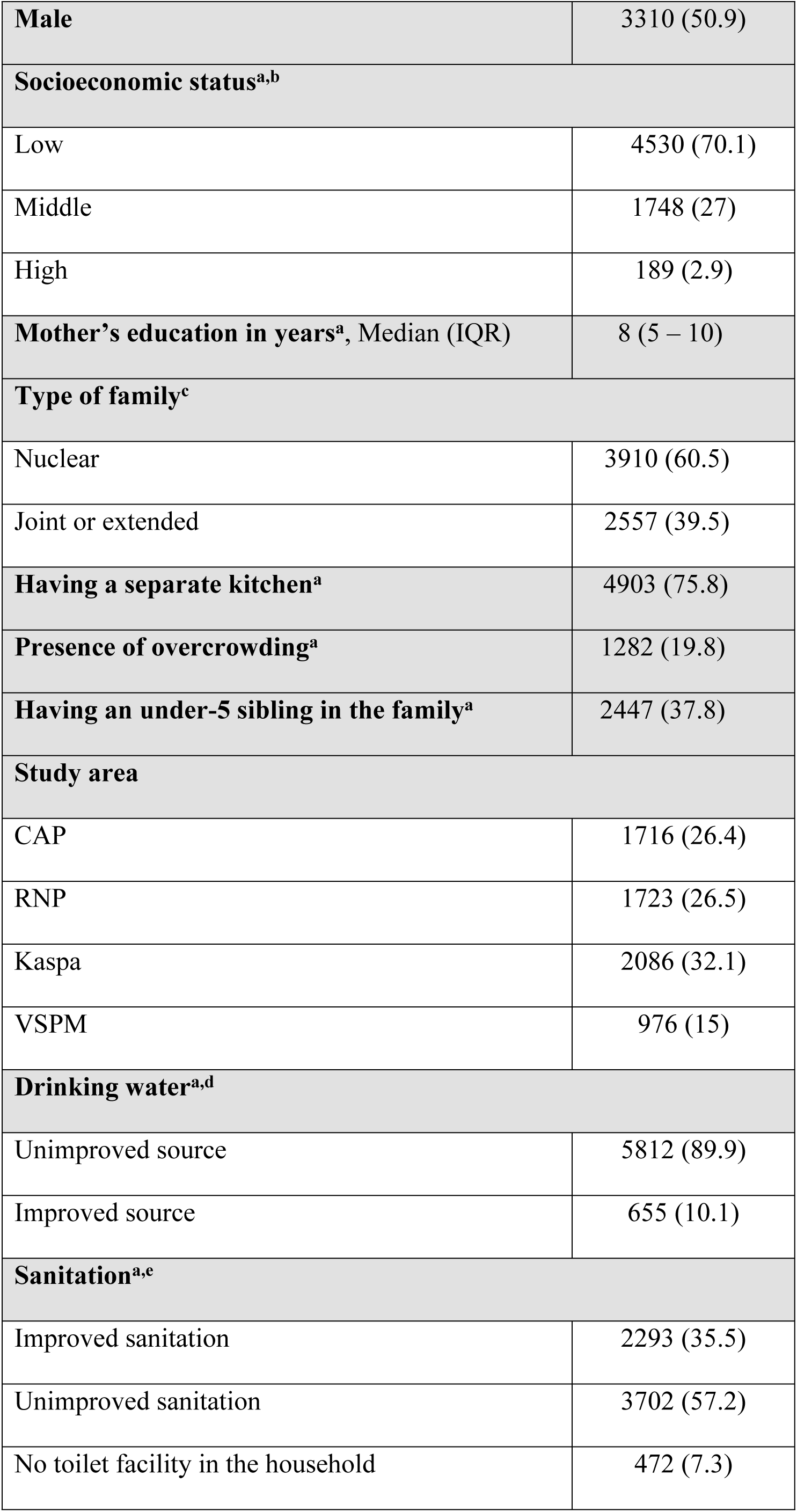
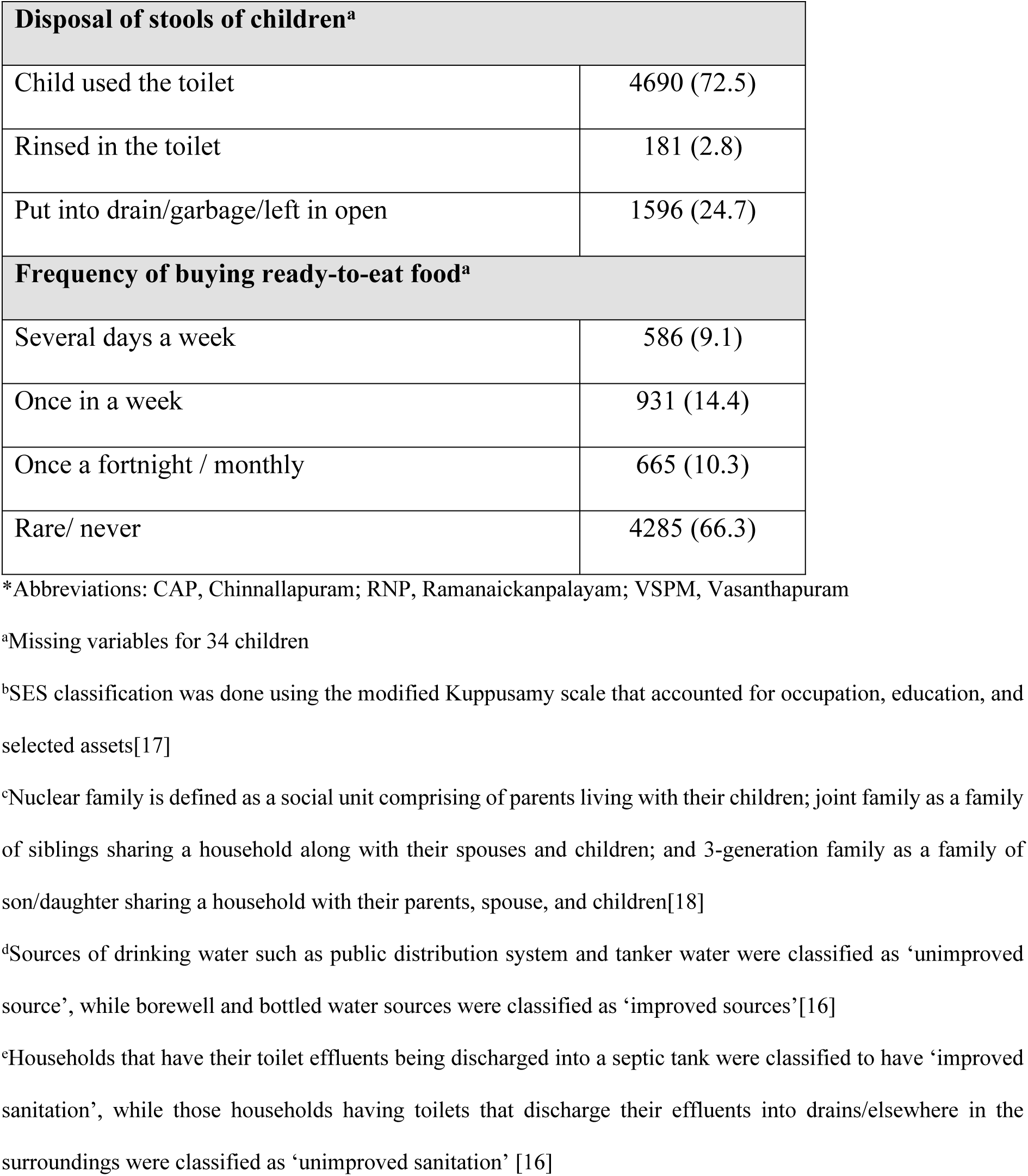
Sociodemographic and WaSH characteristics of children in the Surveillance for Enteric fever in India cohort, 2017-19 (N=6501)

| Characteristics | N (%) |
| --- | --- |
| **Age category at recruitment (years)** |  |
| 0.5 - < 2 | 842 (13) |
| 2 - <5 | 1676 (25.8) |
| 5 - 15 | 3983 (61.3) |
| **Male** | 3310 (50.9) |
| **Socioeconomic status<sup>a,b</sup>** |  |
| Low | 4530 (70.1) |
| Middle | 1748 (27) |
| High | 189 (2.9) |
| **Mother's education in years<sup>a</sup>, Median (IQR)** | 8 (5 – 10) |
| **Type of family<sup>c</sup>** |  |
| Nuclear | 3910 (60.5) |
| Joint or extended | 2557 (39.5) |
| **Having a separate kitchen<sup>a</sup>** | 4903 (75.8) |
| **Presence of overcrowding<sup>a</sup>** | 1282 (19.8) |
| **Having an under-5 sibling in the family<sup>a</sup>** | 2447 (37.8) |
| **Study area** |  |
| CAP | 1716 (26.4) |
| RNP | 1723 (26.5) |
| Kaspa | 2086 (32.1) |
| VSPM | 976 (15) |
| **Drinking water<sup>a,d</sup>** |  |
| Unimproved source | 5812 (89.9) |
| Improved source | 655 (10.1) |
| **Sanitation<sup>a,e</sup>** |  |
| Improved sanitation | 2293 (35.5) |
| Unimproved sanitation | 3702 (57.2) |
| No toilet facility in the household | 472 (7.3) |

| **Disposal of stools of children<sup>a</sup>** |  |
| --- | --- |
| Child used the toilet | 4690 (72.5) |
| Rinsed in the toilet | 181 (2.8) |
| Put into drain/garbage/left in open | 1596 (24.7) |
| **Frequency of buying ready-to-eat food<sup>a</sup>** |  |
| Several days a week | 586 (9.1) |
| Once in a week | 931 (14.4) |
| Once a fortnight / monthly | 665 (10.3) |
| Rare/ never | 4285 (66.3) |

\*Abbreviations: CAP, Chinnallapuram; RNP, Ramanaickanpalayam; VSPM, Vasanthapuram
<sup>a</sup>Missing variables for 34 children
<sup>b</sup>SES classification was done using the modified Kuppasamy scale that accounted for occupation, education, and selected assets[17]
<sup>c</sup>Nuclear family is defined as a social unit comprising of parents living with their children; joint family as a family of siblings sharing a household along with their spouses and children; and 3-generation family as a family of son/daughter sharing a household with their parents, spouse, and children[18]
<sup>d</sup>Sources of drinking water such as public distribution system and tanker water were classified as ‘unimproved source’, while borewell and bottled water sources were classified as ‘improved sources’[16]
<sup>e</sup>Households that have their toilet effluents being discharged into a septic tank were classified to have ‘improved sanitation’, while those households having toilets that discharge their effluents into drains/elsewhere in the surroundings were classified as ‘unimproved sanitation’ [16]

Between November 1, 2017, and October 31, 2019, 6501 children were followed up, and 3612 diarrhoeal episodes were reported during 11600.7 CYO. The overall incidence rate of diarrhoea was estimated as 31.1 per 100 CYO, with children aged 6 months to 5 years reporting a higher incidence rate of 58.6 episodes per 100 child years (**Table 1**). With regard to seasonal trends in reporting diarrhoea cases, months between June and August in 2018 and 2019 reported a higher number of cases, coinciding with the monsoon season in Vellore (**Figure 1**).

**Fig 1:**
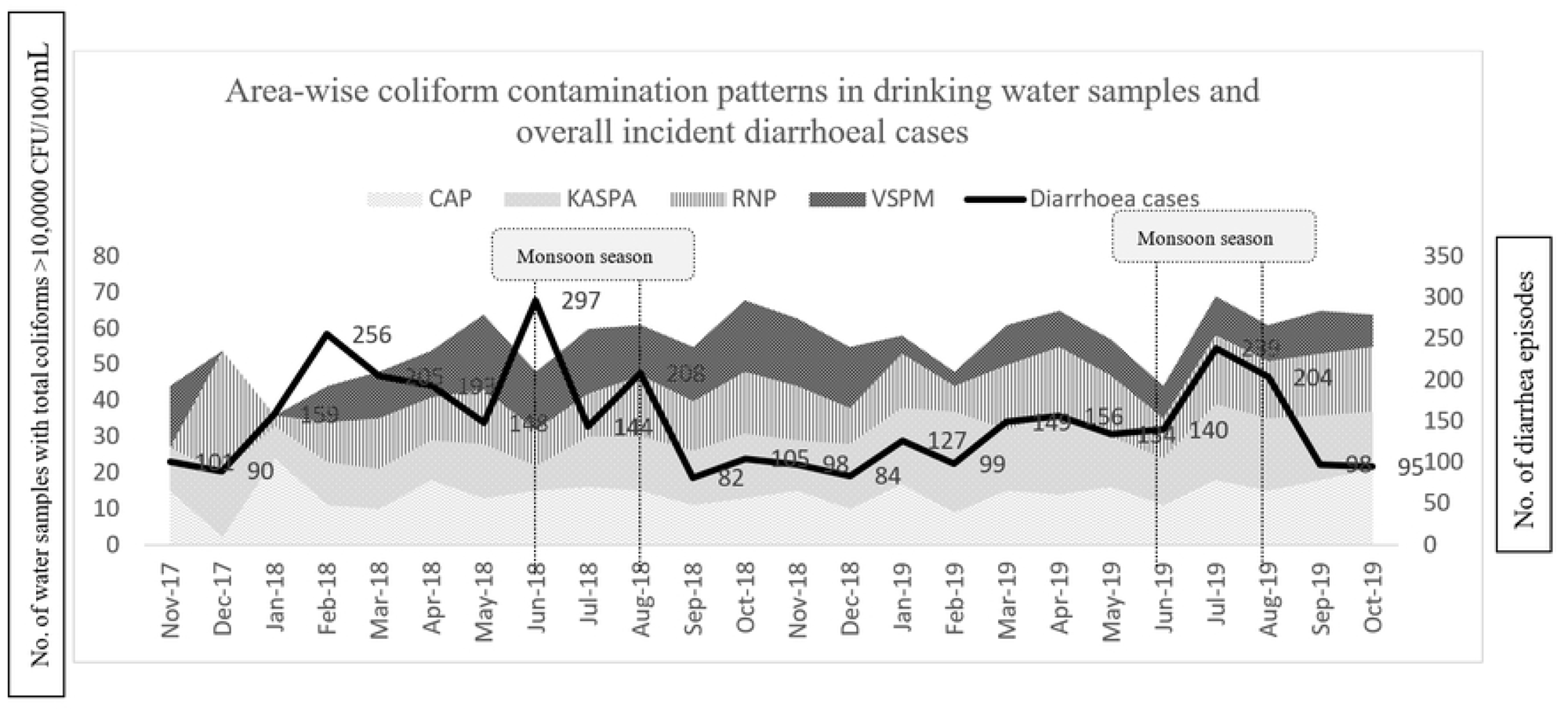
Seasonal trend of diarrhoea in children and coliform contamination of household drinking water in the SEFI cohort, Vellore 2017-2019

Assessment of the quality of drinking water samples in the study area during the surveillance period showed that 1346 (74.6%) out of 1804 drinking water samples collected from study households had coliform counts greater than 10,000/100 ml. Only 110 (6.1%) samples showed a residual chlorine level of 2 PPM *(data not shown)*. Area-wise analysis showed that contamination with coliform counts >10,000 CFU/100 mL was found to be highest in VSPM, followed by RNP and Kaspa, with CAP recording the least number of samples with coliform contamination (**Figure 1**)

Children in the younger age group of 0.5-2 years [IRR 3.5 (95% CI: 3.2 – 3.8)], residing in areas with denser population such as RNP [IRR: 2.6 (95% CI: 2.3 – 2.8)] and Kaspa [IRR: 1.8 (95% CI: 1.6 – 2.0)] and those households with unimproved sources for drinking water such as public distribution system and water supplied by mobile tankers were at the highest risk for diarrhoea. With regard to sanitation at the household level, there was no significant increase in the incidence of diarrhoea among households that had their toilet effluents let into open drains/elsewhere (unimproved sanitation) compared to those that left their effluents in the septic tanks (improved sanitation). In the case of hygiene practices at the household level, unhygienic practices such as rinsing children’s stools in toilets [IRR 1.3 (95% CI: 1.0 – 1.5)] or disposing of into drains/garbage [IRR 1.3 (95% CI: 1.2 – 1.4)] had higher chances of reporting diarrhoeal cases than households where children used toilets. Similarly, households with a higher frequency of buying ready-to-eat food several days a week [IRR 1.2 (95% CI: 1.0 – 1.3)] had a higher risk for diarrhoea compared to those who rarely bought ready-to-eat food **(Table 3).**

**Table 3:** Factors associated with incidence of diarrhoeal episodes among children 6 months – 15 years in Surveillance for Enteric Fever in India cohort, 2017-19 (N=6400)

| Characteristics | Unadjusted<br>IRR<br>with 95% CI | Adjusted IRR<br>with 95% CI |
| --- | --- | --- |

| Age category at recruitment (years) |  |  |
| --- | --- | --- |
| 0.5 - < 2 | **3.8 (3.5 – 4.1)** | **3.5 (3.2 – 3.8)** |
| 2 - <5 | **1.4 (1.3 – 1.5)** | **1.4 (1.3 – 1.5)** |
| 5 - 15 | *ref* | *ref* |
| Socioeconomic status<sup>a</sup> |  |  |
| Low | **0.7 (0.6 – 0.8)** | 0.9 (0.7 – 1.1) |
| Middle | **0.8 (0.7 – 1.0)** | 0.9 (0.8 – 1.1) |
| High | *ref* | *ref* |
| **Mother's education in years, Median (IQR)** | **1.0 (1.0 - 1.1)** | **1.0 (1.0 – 1.0)** |
| Type of family<sup>b</sup> |  |  |
| Joint or extended | **1.3 (1.2 – 1.4)** | 1.1 (1.0 – 1.1) |
| Nuclear | *ref* | *ref* |
| **Having an under-5 sibling in the family** | **1.1 (1.0 – 1.2)** | **0.9 (0.8 – 0.9)** |
| Study area |  |  |
| CAP | *ref* | *ref* |
| RNP | **2.4 (2.2 – 2.6)** | **2.6 (2.3 – 2.8)** |
| Kaspa | **1.8 (1.6 – 2.0)** | **1.8 (1.6 – 2.0)** |
| VSPM | **1.6 (1.4 – 1.8)** | **1.4 (1.2 – 1.6)** |
| Source of drinking water<sup>c</sup> |  |  |
| Unimproved source | 1.1 (1.0 – 1.2) | **1.2 (1.0 – 1.3)** |
| Improved source | *ref* | *ref* |
| Sewage disposal<sup>d</sup> |  |  |
| Improved sanitation | *ref* | *ref* |
| Unimproved sanitation | 0.9 (0.8 – 1.0) | 1.0 (0.9 – 1.0) |
| No toilet facility in the household | 1.0 (0.8 – 1.1) | 1.0 (0.9 – 1.2) |
| **Disposal of stools of children** |  |  |
| Child used the toilet | *ref* | *ref* |
| Rinsed in the toilet | **1.9 (1.6 – 2.3)** | **1.3 (1.0 – 1.5)** |
| Put into drain/garbage/left in open | **1.6 (1.5 – 1.7)** | **1.3 (1.2 – 1.4)** |
| **Frequency of buying ready-to-eat food** |  |  |
| Several days a week | **1.2 (1.0 – 1.3)** | **1.2 (1.0 – 1.3)** |
| Once in a week | **1.7 (1.1 – 1.3)** | 0.9 (0.8 – 1.0) |
| Once a fortnight / monthly | **1.3 (1.1 – 1.4)** | 1.0 (0.9 – 1.1) |
| Rare/ never | *ref* | *ref* |

\*Abbreviations: CAP, Chinnallapuram; RNP, Ramanaickanpalayam; VSPM, Vasanthapuram
<sup>a</sup>SES classification was done using the modified Kuppusamy scale that accounted for occupation, education, and selected assets[17]
<sup>b</sup>Nuclear family is defined as a social unit comprising of parents living with their children; joint family as a family of siblings sharing a household along with their spouses and children; and 3-generation family as a family of son/daughter sharing a household with their parents, spouse, and children[18]
<sup>c</sup>Sources of drinking water such as public distribution system and tanker water were classified as ‘unimproved source’, while borewell and bottled water sources were classified as ‘improved sources’[16]
<sup>d</sup>Households that have their toilet effluents being discharged into a septic tank were classified to have ‘improved sanitation’, while those households having toilets that discharge their effluents into drains/elsewhere in the surroundings were classified as ‘unimproved sanitation’ [16]

## Discussion

This cohort study assessed the incidence of diarrhoeal episodes among children in an urban settlement of south India and found it to be 31.1 per 100 CYO, with children aged between 6 months and 5 years at a significantly higher risk than those between 5 and 15 years. A clear seasonal trend was observed, with a peak in reported cases during the monsoon months (south-west monsoon between June to August). Risk factors identified to be associated with the incidence of diarrhoeal episodes include younger age (3.5 times higher risk), residence in densely populated areas (1.8 to 2.6 times), use of unimproved drinking water sources such as the public distribution system or water supply by mobile tanker (1.2 times) and poor food hygiene practices (1.2 times).

The estimates of diarrhoeal episode incidence in children of age 0.5-5 years in our study (58.6/100 CYO) is less than the national estimates, which report a higher incidence in rural areas (171 episodes per 100 CYO) than urban (109 episodes per 100 CYO) among children aged 0–6 years [11]. A birth cohort study from Vellore conducted during 2008-2011 reported an incidence of 310 episodes per 100 CYO during infancy and 200 episodes per 100 CYO in the second year of life, which is higher than our estimates of 58.6 per 100 CYO (0.58 per CYO) in 0.5–5 years [19]. The relatively lower estimates in our study compared to the previous study from Vellore conducted almost a decade before, could be due to the improvement in water and sanitation conditions between the time frame of these studies [19]. Also, as Sarkar et al. show, a greater incidence in the younger age group could be due to greater susceptibility to diarrhoeal pathogens in these children compared to older children [19]. Other community-based cohort studies conducted in Sevagram (2010–11) and urban Goa reported diarrheal incidence rates of 65 per 100 CYO and 12.4 per 100 CYO among children under five, highlighting the heterogeneity in estimates from different study settings [5,20]. This heterogeneity in estimates may be due to study-level differences with respect to diverse risk settings, period of study and age groups considered in the studies.

A distinct seasonal variation in diarrheal disease incidence was observed in our study, with a peak in cases during the south-west monsoon season. This pattern aligns with the results of a previous research that observed a peak in diarrhoeal deaths during monsoon seasons in India [9]. Similar seasonal trends have been documented in a study conducted in Nepal over a 13-year period (2002–2014), where diarrheal disease incidence increased by 21% during the monsoon season [21]. Additionally, a 2018 time-series analysis from Delhi provides further evidence supporting the seasonal increase in diarrheal cases and suggesting monsoon-related flooding, and poor sewage and drainage infrastructure potentially contribute to water contamination, subsequently leading to diarrhoeal outbreaks [22] Heavy rainfall often overwhelms sewage systems, causing contamination of drinking water sources with faecal matter. Additionally, higher temperatures and rainfall during the monsoon season create an environment conducive to the proliferation of diarrheal pathogens, increasing transmission risk [23].

Our study found that approximately 75% of household water samples were contaminated with coliforms, indicating a significant risk of waterborne disease transmission. This finding is consistent with a study conducted in the Vellore district in 2017, which reported that 60% of household water samples were faecal contaminated [24]. The presence of coliforms in drinking water is a well-established risk factor for diarrheal diseases, as it indicates probable contamination with human or animal faeces, implying contamination with pathogenic bacteria, viruses, and protozoa. Studies have demonstrated a direct correlation between faecal-contaminated drinking water and an increased incidence of diarrheal diseases. For instance, a study conducted in rural Bangladesh in 2015 found that households with faecally contaminated drinking water had significantly higher rates of diarrheal illnesses [25]. Additionally, a 2018 systematic review and meta-analysis further supported this association, highlighting the role of unsafe drinking water as a major contributor to the global burden of diarrheal diseases [26].

Environmental risk factors played a significant role in the diarrheal disease burden observed in our study. Unimproved sanitation practices, particularly the unhygienic disposal of children’s excreta, was found to be significant contributor to diarrheal disease in young children, and this aligns with a cross-sectional survey conducted in rural Odisha (2021), which identified that unsafe management of human excreta posed a 2.4 times higher risk for diarrhoeal episodes among children aged 6 months to 1 year [27]. Additionally, a study from Ethiopia (2016–2021) supports our findings, demonstrating a strong association between the availability of toilet facilities at the household and diarrheal disease prevalence [28]. The lack of safely managed sanitation facilities results in the widespread contamination of the water bodies that supply drinking water to households with enteric pathogens, facilitating feco-oral transmission through water, food, and/or direct person-to-person contact. Thus, children residing in areas with unimproved sanitation and densely populated urban settlements, as in this study, are at higher risk for water contamination and suffer from higher diarrhoeal illness episodes. In addition to WaSH factors, frequent consumption of ready-to-eat foods was associated with higher diarrhoeal incidence in our study, likely due to poor food hygiene—an observation consistent with findings from Bangladesh, highlighting food contamination as a major risk in low-resource settings [29]. In line with Sustainable Development Goal 6, attaining optimal WaSH facilities in urban areas is required to bring down diarrhoeal burden at the population level.

A notable strength of our study is the intensive surveillance data from the longitudinal SEFI cohort of children aged <15 years, providing a comprehensive understanding of the diarrheal disease burden. Most previous studies have primarily focused on children under five, while this study provides stratified estimates for children aged 0.5 –5 years and 5–15 years, thereby providing estimates on the diarrheal burden in older children as well. However, our study has certain limitations. Although our study captured diarrheal events in the past week, information on the start and end dates of these episodes was limited. Exclusion of children under 6 months, as per the protocol of the original study estimating enteric fever incidence, resulted in the lack of data from this age group, possibly underestimating the overall burden.

## Conclusion

This cohort study assessed the incidence of diarrhoeal episodes among children in an urban settlement in south India as 31.1 per 100 child-years of observation, with children aged between 6 months and 5 years at higher risk for diarrhoeal diseases. Approximately 8 in 10 children in urban Vellore do not have safe access to drinking water, putting them, especially the under-5 children, at a high risk for diarrheal illnesses. Improvement in quality parameters of WaSH, and not mere geographic expansion, is pivotal in reducing the burden of diarrheal diseases in children.

## Data Availability

Data will be provided upon request to the corresponding author.

## Acknowledgement

We thank the parents and children of the SEFI study, as well as the field research assistants and field supervisors (Mr Prabhu, Mr Selvakumar, and Mr Raja) of the Vellore SEFI cohort. We thank Mr Ravi for collecting drinking water samples from study households. We are thankful to Mrs Sheela and her team at the Wellcome Trust Research Lab for water sample analysis. We are grateful for the support of the study clinic team and the study physicians for the clinical support.

